# Trends in Complications among Patients undergoing Aortic Valve Replacement (AVR) in the United States

**DOI:** 10.1101/2023.06.27.23291980

**Authors:** James E. Harvey, Samir R. Kapadia, David J. Cohen, Ankur Kalra, William Irish, Candace Gunnarsson, Michael Ryan, Soumya Chikermane, Christin Thompson, Rishi Puri

## Abstract

**Background:** The treatment of severe aortic stenosis has evolved considerably since the introduction of transcatheter aortic valve replacement (TAVR), yet trends in in-hospital complications for patients undergoing TAVR or surgical aortic valve replacement (SAVR) at a national level have yet to be evaluated.

**Methods:** We performed a retrospective cohort study using Medicare data to evaluate temporal trends in complications among beneficiaries aged ≥65 years treated with elective isolated transfemoral TAVR or SAVR between 2012 and 2019. The study endpoint was the occurrence of a major complication during the index AVR hospitalization, defined as a composite outcome. Multivariable logistic regression was used to assess odds of complications for TAVR and SAVR, individually over time, after adjusting for baseline characteristics. Another risk-adjusted model assessed the risk of complications for TAVR vs SAVR, over time.

**Results:** The cohort included 211,212 patients (mean age:78.6±7.3 years, female:45.0%). Complication rates following elective isolated AVR decreased from 49% in 2012 to 22% in 2019. These reductions were more pronounced for TAVR (41%->19%, delta=22%) than SAVR (51%->47%, delta=4%). After risk adjustment, the risk of any complication with TAVR was 47% (p<0.0001) lower compared to SAVR in 2012, and 78% (p<0.0001) lower in 2019. TAVR was independently associated with reduced odds of complications each year compared to 2012, with the magnitude of benefit increasing over time (2013 vs 2012: OR=0.89(0.81-0.97); 2019 vs 2012: OR=0.35(0.33-0.38)).

**Conclusions:** Between 2012-2019, the risk of complications after AVR among Medicare beneficiaries decreased significantly, with larger absolute and relative changes among patients treated with TAVR than SAVR.

## INTRODUCTION

Historically, surgical aortic valve replacement (SAVR) has been the standard treatment for patients with aortic stenosis (AS). However, the introduction of transcatheter aortic valve replacement (TAVR) has profoundly changed the management of patients with AS, and TAVR has now surpassed SAVR in terms of annual procedural volume in United States (US). ^1-4^

While both treatment modalities remain fundamental therapeutic approaches to patients with AS, over the past decade, there have been considerable improvements in TAVR including advances in procedural technique, device technologies, and patient selection. In contrast, the procedural and technological aspects of SAVR have remained more constant given that it is a more mature procedure with a well-established track record and historically excellent outcomes. ^5^ Although previous studies have explored the impact of treatment evolution on short and long-term mortality, no studies have focused on the changes in complications between TAVR and SAVR. ^2,6-8^ In particular, it is unclear how changes in these factors (including patient profile) have impacted major procedural complications over time and across different forms of AVR.

Thus, the purpose of the present analysis was to assess temporal trends in in-hospital complication rates among Medicare beneficiaries receiving aortic valve replacement (AVR) over time (2012-2019) in a real-world setting reflective of US practice, and to examine whether these trends differed between patients undergoing TAVR or SAVR.

## METHODS

### Data Source

This retrospective cohort study used Medicare fee for service (FFS) data. The Medicare FFS payer data includes information on healthcare services that are covered for beneficiaries enrolled in Medicare Parts A and B. All data used to perform this analysis were de-identified and accessed in compliance with the Health Insurance Portability and Accountability Act. As a retrospective analysis of a de-identified database, the research was exempt from IRB review under 45 CFR 46.101(b), and the need for individual patient consent was waived.

### Population of Interest

The analytic cohort for our study included Medicare FFS beneficiaries who underwent elective isolated AVR (transfemoral TAVR (TF-TAVR), or SAVR) between 2012 and 2019 and who were aged 65 years or older at the time of their procedure. The earliest AVR admission during the study period was considered the index AVR. For this study, the admission status variable in Medicare was used to define urgent/emergent or elective AVRs. Patients also had to have at least 6 months of continuous enrollment in Medicare Parts A and B prior to AVR in order to allow for identification of comorbidities. Patients were excluded if they underwent concomitant aortic root repair, coronary artery bypass grafting (CABG), additional valve procedures, or had a diagnosis of endocarditis.

### Study Endpoints

The primary study endpoint was a composite of major in-hospital complications including death, new atrial fibrillation, acute kidney injury (AKI), acute myocardial infarction (AMI), aortic rupture, hemorrhage, post-operative infection, pacemaker, prolonged ventilation, respiratory failure, septicemia, stroke, or vascular complications.

### Covariates

Covariates were obtained from Medicare claims and included patient demographics (age, sex, race, and region) and comorbid conditions included in the Elixhauser Comorbidity Index (ECI). ^9^ Previous studies have shown that the ECI is associated with in-hospital mortality, length of stay, and adverse events across a broad range of medical conditions.

### Statistical Analyses

Analyses were performed in the elective isolated AVR cohort as well as stratified analysis in the elective TF-TAVR, and elective isolated SAVR sub-cohorts, individually, to assess the odds of in-hospital complications over time.

Patient demographics and ECI were summarized for the overall AVR population as well as for the TAVR and SAVR subgroups. Continuous variables are reported as mean and standard deviation, and categorical variables are reported as frequencies or percentages. Statistical models were run using SAS 9.4. Multivariable logistic regression models were developed to assess the odds of complications for TAVR vs SAVR during index hospitalization (primary exposure of interest: type of AVR) and were adjusted for baseline differences in patient age, sex, region and ECI Index. Calendar year was interacted with each covariate and when significant, odds ratios are reported for each year separately. A 2-sided p-value of <0.05 was considered statistically significant. Odds ratios and 95% confidence intervals (CIs) are provided as measures of strength of association and precision, respectively.

In addition to this, stratified analysis by elective TAVR and elective isolated SAVR was performed to assess the risk of complications over time (primary exposure of interest: Index Year (ref=2012)) in the TAVR and SAVR groups, respectively.

## RESULTS

A total of 429,967 Medicare beneficiaries underwent AVR between 2012 and 2019. Of these, 211,212 met criteria for inclusion in the study (*Figure 1*) of which 69.2% were TAVR (n= 146,239) and 30.8% were SAVR (n= 64,973) of the AVRs. *Figure 2* displays the distribution of elective isolated SAVR and elective TF-TAVR by the index year. The total number of elective isolated AVRs performed annually increased from 13,310 in 2012 (25 AVRs per 100,000 Medicare FFS beneficiaries) to 40,015 in 2019 (62 AVRs per 100,000 Medicare FFS beneficiaries). The proportion of AVRs that were performed surgically decreased from 75% in 2012 to 10% in 2019.

**Figure 1:**
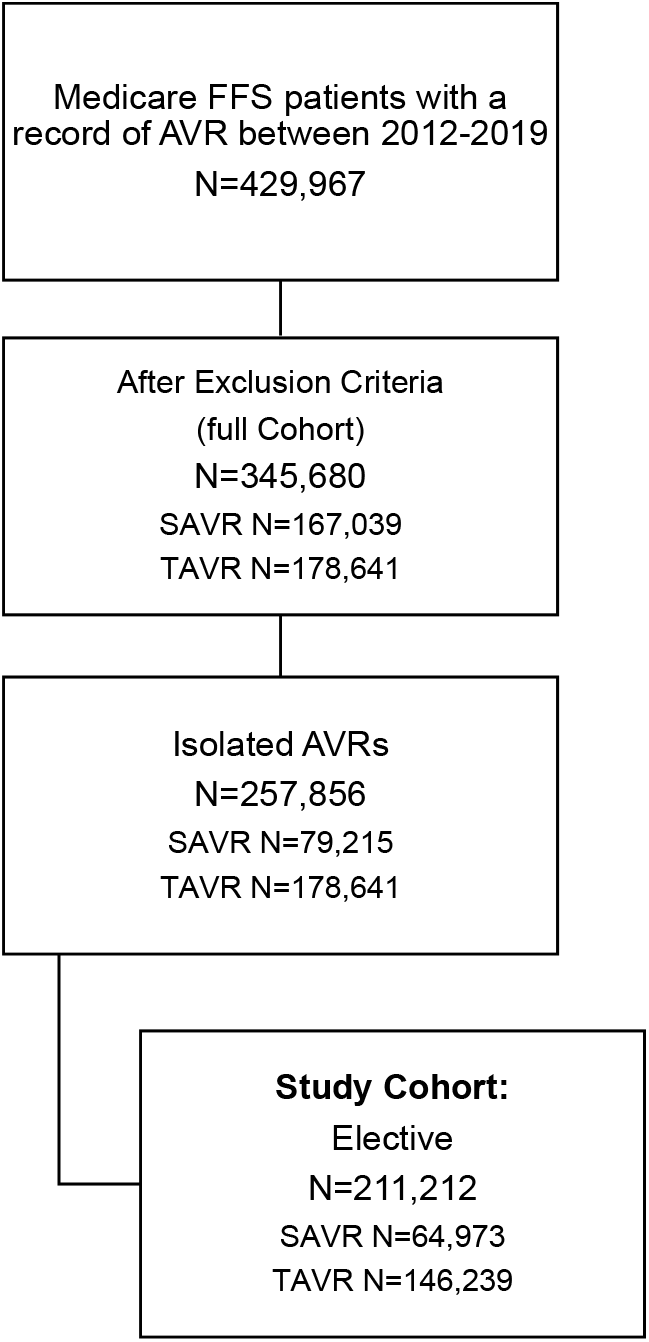
Attrition Diagram.

**Figure 2:**
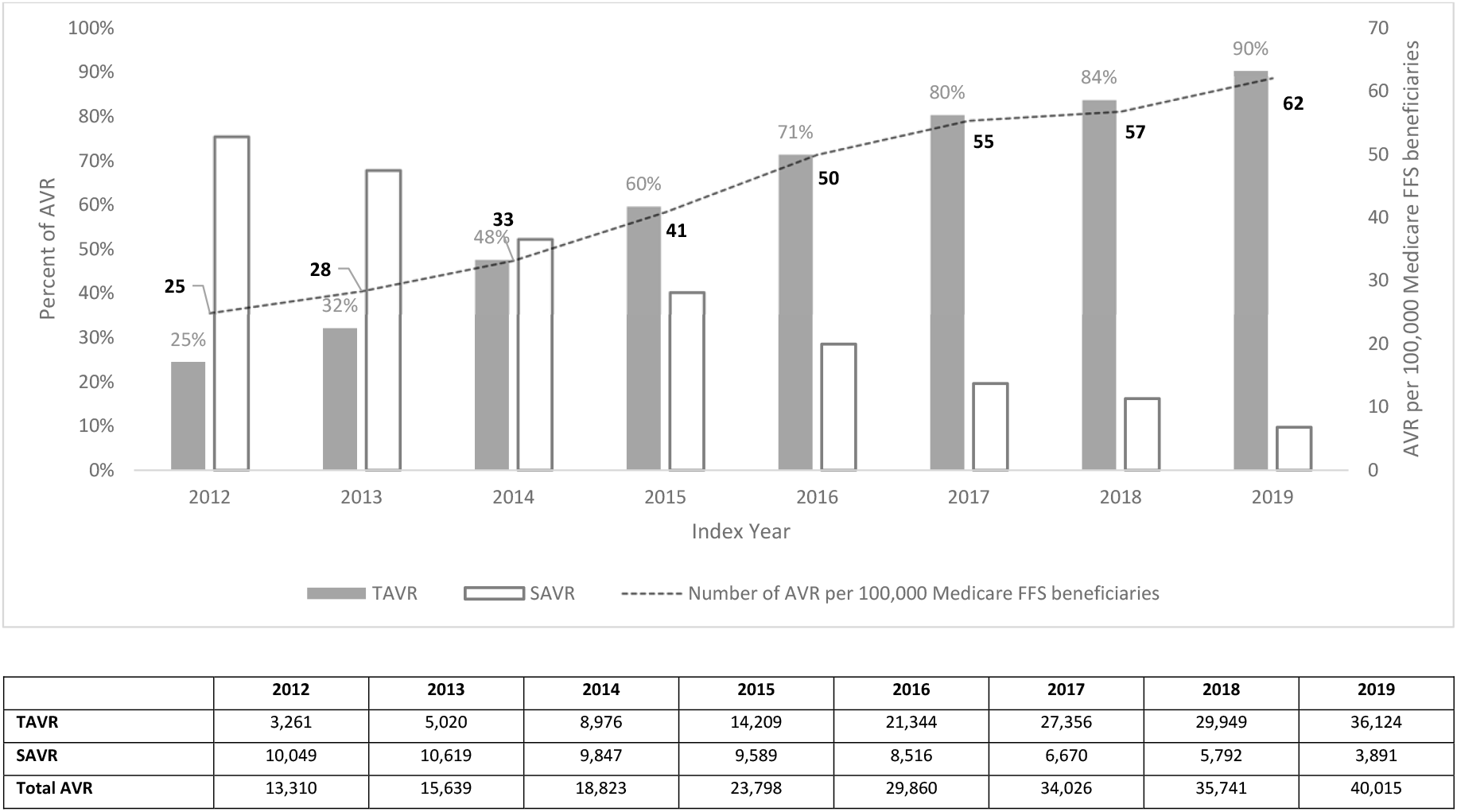
Proportion of TAVR and SAVR in the elective isolated AVR Cohort, and the number of AVRs per 100,000 Medicare FFS beneficiaries by Index Year.

### Baseline characteristics

*Table 1* displays baseline patient characteristics for each cohort. In the cohort of 211,212 AVR patients, the mean age was 80.6±6.9 years for TAVR and 74.0±6.0 years for SAVR. The majority of patients in both the TAVR and SAVR cohorts were male (TAVR: 53.9%, SAVR: 57.5%), and Caucasian (TAVR: 93.7%, SAVR: 92.9%). The mean ECI in the TAVR and SAVR groups were 6.4±3.5 and 4.4±2.7 respectively.

**Table 1:**
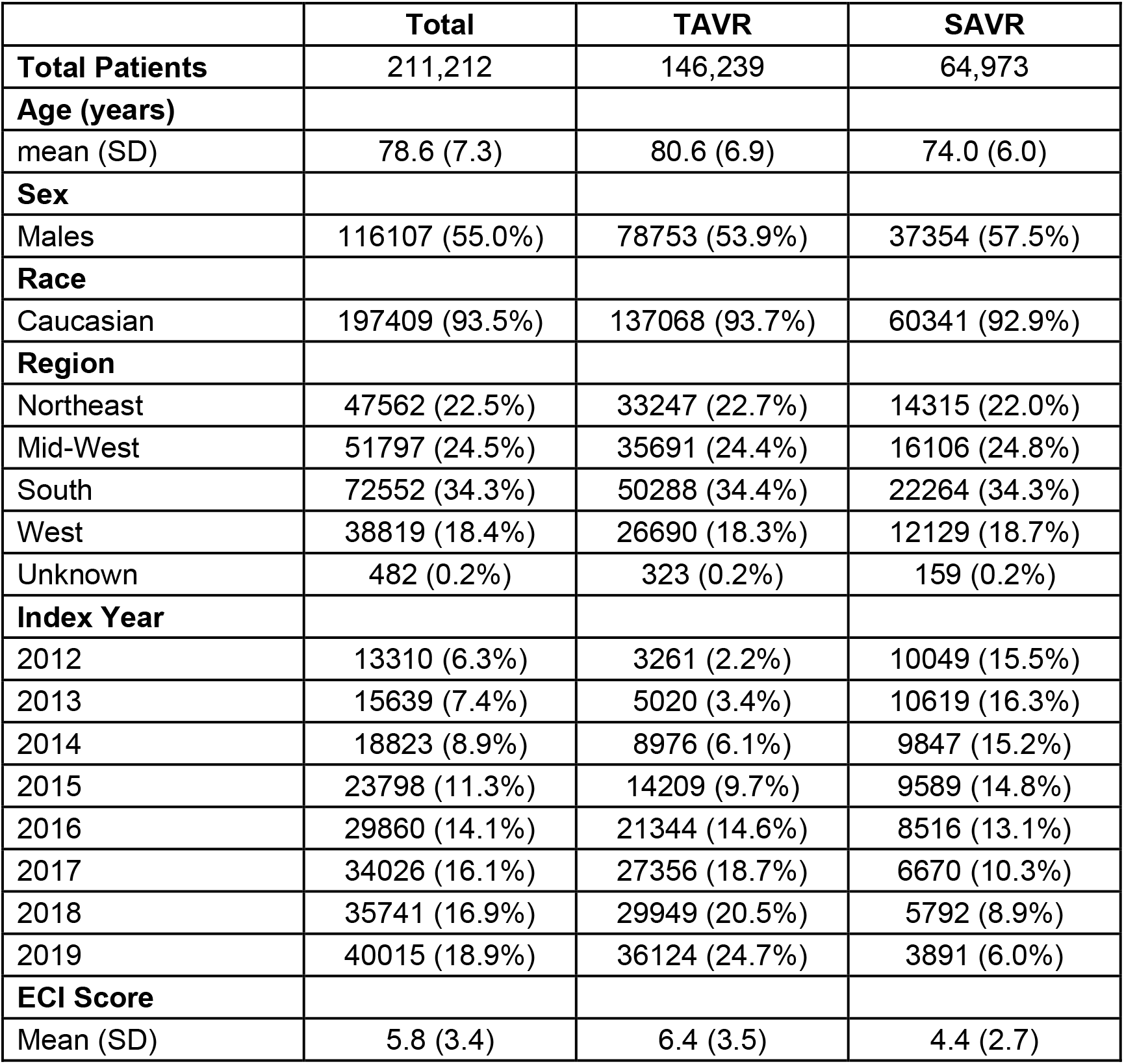
Patient Characteristics of the elective isolated AVR Cohort.

|  | **Total** | **TAVR** | **SAVR** |
| --- | --- | --- | --- |
| **Total Patients** | 211,212 | 146,239 | 64,973 |
| **Age (years)** |  |  |  |
| mean (SD) | 78.6 (7.3) | 80.6 (6.9) | 74.0 (6.0) |
| **Sex** |  |  |  |
| Males | 116107 (55.0%) | 78753 (53.9%) | 37354 (57.5%) |
| **Race** |  |  |  |
| Caucasian | 197409 (93.5%) | 137068 (93.7%) | 60341 (92.9%) |
| **Region** |  |  |  |
| Northeast | 47562 (22.5%) | 33247 (22.7%) | 14315 (22.0%) |
| Mid-West | 51797 (24.5%) | 35691 (24.4%) | 16106 (24.8%) |
| South | 72552 (34.3%) | 50288 (34.4%) | 22264 (34.3%) |
| West | 38819 (18.4%) | 26690 (18.3%) | 12129 (18.7%) |
| Unknown | 482 (0.2%) | 323 (0.2%) | 159 (0.2%) |
| **Index Year** |  |  |  |
| 2012 | 13310 (6.3%) | 3261 (2.2%) | 10049 (15.5%) |
| 2013 | 15639 (7.4%) | 5020 (3.4%) | 10619 (16.3%) |
| 2014 | 18823 (8.9%) | 8976 (6.1%) | 9847 (15.2%) |
| 2015 | 23798 (11.3%) | 14209 (9.7%) | 9589 (14.8%) |
| 2016 | 29860 (14.1%) | 21344 (14.6%) | 8516 (13.1%) |
| 2017 | 34026 (16.1%) | 27356 (18.7%) | 6670 (10.3%) |
| 2018 | 35741 (16.9%) | 29949 (20.5%) | 5792 (8.9%) |
| 2019 | 40015 (18.9%) | 36124 (24.7%) | 3891 (6.0%) |
| **ECI Score** |  |  |  |
| Mean (SD) | 5.8 (3.4) | 6.4 (3.5) | 4.4 (2.7) |

### Trends in complications for TAVR and SAVR

Figure 3 displays the unadjusted rates for the primary endpoint over time, by approach (SAVR versus TAVR). The complication rates following AVR decreased every year from 49% in 2012 to 22% in 2019, and the TAVR complication rate was consistently lower than that of SAVR for all years studied (41% vs. 51% in 2012 and 19% vs. 47% in 2019, respectively). Additionally, the decrease in complication rate between 2012 and 2019 was far more pronounced for TAVR (41% to 19%, delta = 22%) than for SAVR (51% to 47%, delta = 4%). Appendix Figure S1 (A-M) summarizes the crude percentages of individual complications over time for the elective isolated TAVR and SAVR groups.

**Figure 3:**
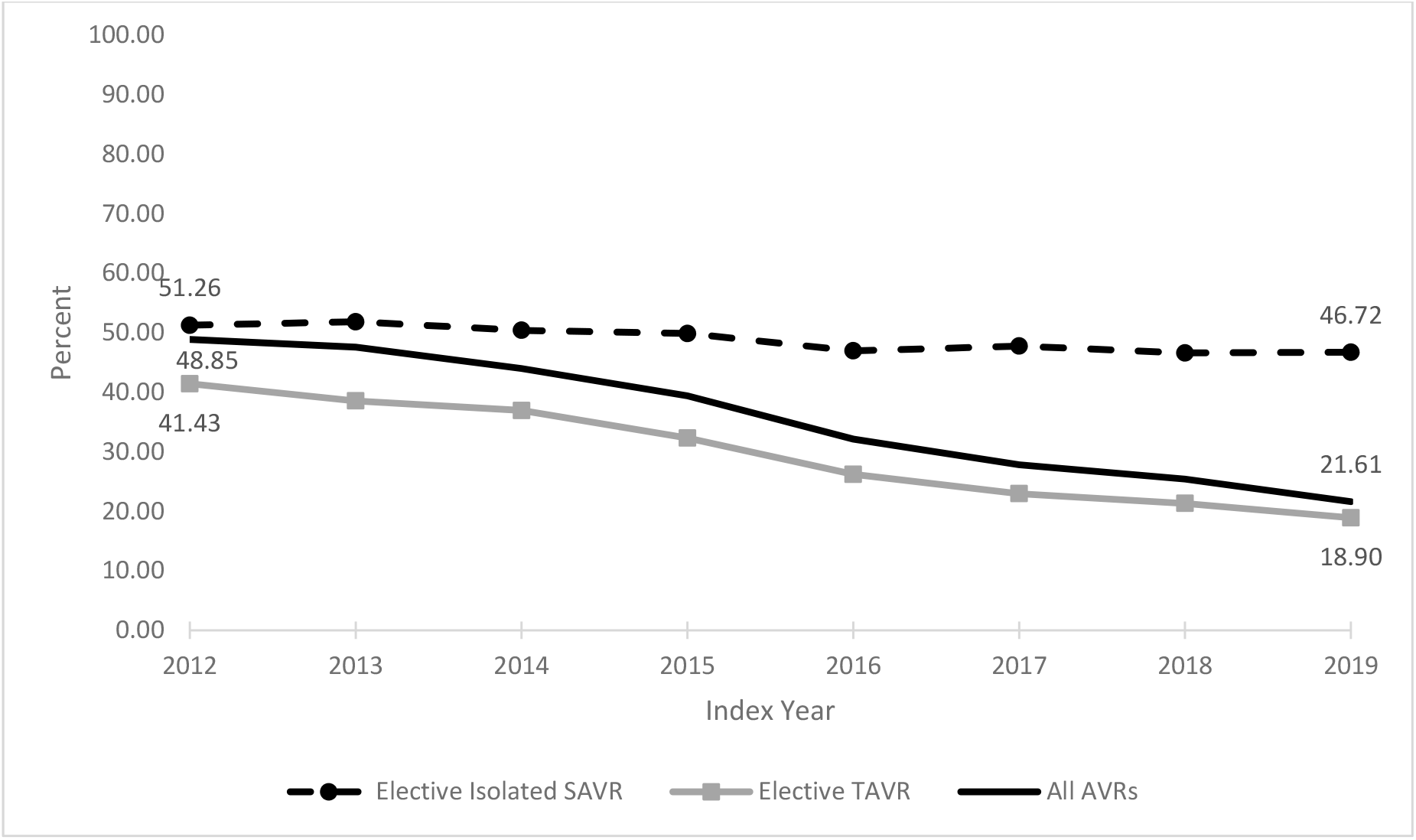
Unadjusted composite complication rates over time for the elective isolated AVR Cohort, elective TAVR and elective isolated SAVR.

The most common complications among TAVR patients in 2019 were pacemaker implantation, acute kidney injury, hemorrhage, and new atrial fibrillation. The most common complications among SAVR patients in 2019 were new atrial fibrillation, acute kidney injury, respiratory failure, and pacemaker implantation.

### Multivariable models to assess odds of complications during hospitalization

Figures 4A and 4B present the multivariable logistic regression results for the elective TF-TAVR and elective isolated SAVR groups by index year, relative to index year 2012. In the TAVR subgroup, the adjusted odds of the composite endpoint decreased progressively over time from 0.89 (95% CI 0.81-0.97) in 2013 to 0.35 (95% CI 0.33-0.38) in 2019. In the SAVR subgroup, the adjusted odds of the composite endpoint decreased over time as well, although the magnitude of the decrease was less than for TAVR (adjusted OR 1.03 [95% CI 0.98-1.10] in 2013 and 0.91 [95% CI 0.85-0.98] in 2019). When TAVR was compared directly with SAVR, the adjusted odds of complications was consistently lower with TAVR than SAVR over time and decreased from 0.53 (95% CI 0.49-0.58) in 2013 to 0.22 (95% CI 0.22-0.24) in 2019 (Figure 4C).

**Figure 4:**
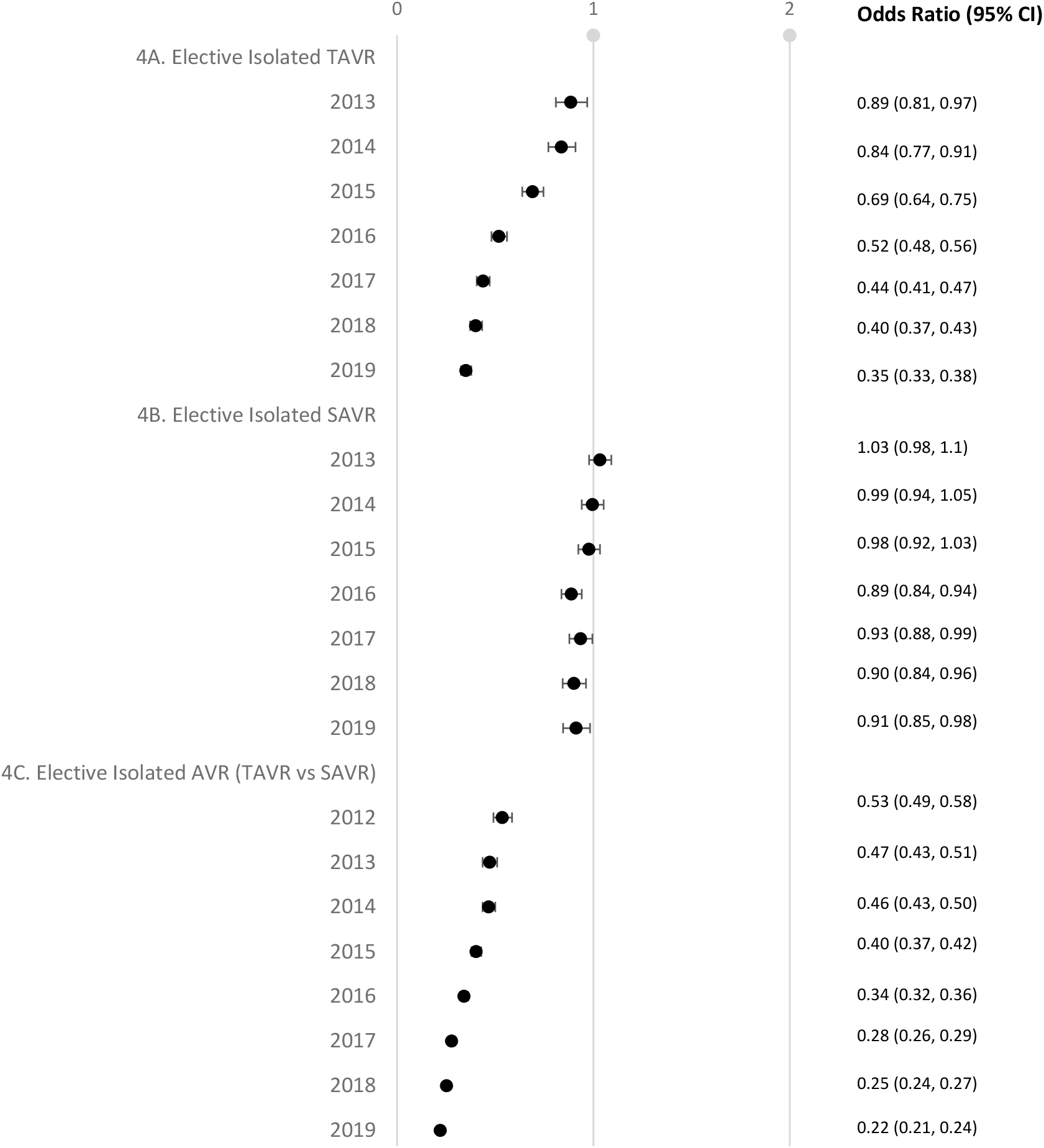
Multivariable model assessing the risk of complications in the elective TAVR cohort (reference: 2012), elective isolated SAVR cohort (reference: 2012), and elective isolated AVR cohort (TAVR vs SAVR).

## Discussion

The present analysis evaluated trends in complication rates of isolated elective AVR in the US over an 8-year period (2012-2019) using the Medicare FFS payer database, which captures the majority of SAVR and TAVR recipients during this time frame. The chosen time period reflects both the commercial inception of TAVR, the introduction of new generations of TAVR devices, as well as the period leading up to and including its use in treating low surgical-risk patients. The pertinent findings are (i) the number of elective isolated AVRs rose significantly over time, increasing from nearly 13,310 (25 patients with AVR per 100,000 Medicare beneficiaries) in 2012 to just under 40,015 (62 patients with AVR per 100,000 Medicare beneficiaries) in 2019, with a fundamental switch in treatment modality from a SAVR/TAVR ratio of 75% / 25% in 2012 to a 10% / 90% equivalent ratio in 2019, (ii) AVR complication rates declined over time, (iii) TAVR-related complications were lower than SAVR across all time periods, and demonstrated a greater reduction (delta -22% vs. -4% for TAVR vs. SAVR-related decline in complications respectively) over time, than SAVR-related complications. These observations illustrate the relatively rapid evolution of TAVR from an immature transcatheter therapy in 2012, to the dominant form of AVR in 2019, enabling a greater proportion of severe AS patients to receive life-saving AVR treatment.

Since its commercial launch in the USA following the results of the PARTNER 1A and 1B trials, a renewed focus and awareness of severe AS, its mortal prognosis, and availability of a non-surgical option has resulted in a significantly higher number of patients with severe AS undergoing AVR, either via a transcatheter approach or surgically. ^10,11^ The culmination of the ability to treat the previously ‘untreatable’ patients deemed unfit/too high risk for SAVR, refinements in patient selection and procedural technique, innovations in transcatheter aortic valve design along with computed tomographic sizing algorithms have all contributed to TAVR emerging as the dominant means of AVR in the US. ^12^

The introduction of TAVR has enabled clinicians to optimize patient selection in terms of preferred AVR technique based upon the patient’s underlying anatomy. For example, patients harboring porcelain aortas, hostile chest re-entry, or poor pulmonary reserve would be considered more suitable for TAVR than for SAVR. ^1^ Conversely, patients with excessive LVOT calcification, small annuli, or low-lying coronary arteries would be more suitable for SAVR than TAVR. ^1^ Additionally, mandatory reporting of TAVR and SAVR-related outcomes to national data repositories has helped to drive improvements in techniques and outcomes across the US. ^13^

Our findings add to existing evidence around TAVR and SAVR outcomes in several ways. A study by Mori et al., has observed similar trends across TAVR and SAVR in terms of mortality, length of stay, and readmission. ^2^ However, this study has not looked at the trends in complications between TAVR and SAVR over time. Lauck et al., assessed trends in the risk of 30-day mortality and readmission from 2012-2019 between TAVR and SAVR found that the rate of these outcomes in TAVR were reducing compared to SAVR until 2016, after which the magnitude of decline started reducing. ^8^ A TVT registry study by Sherwood et al., focused on TAVRs performed between 2011 and 2016, assessed the rates of vascular complications and in-hospital bleeding and found that vascular complications and in-hospital bleeding event rates post-TAVR have declined over time. ^7^ This study however did not assess the contemporary rates of these complications with TAVR, other potential complications of interest for TAVR were not included (for example, atrial fibrillation, pacemaker use, aortic rupture, etc) and there was no comparison with complication rates in SAVR. A study by Arora et al., conducted using the National Inpatient Sample assessed the risk of individual in-hospital complications between TAVR and SAVRs performed between 2012 and 2015 found that compared with SAVR, TAVR was associated with significantly lower incidences of stroke, cardiogenic shock, acute kidney injury, and blood transfusion, but an increased risk of pacemaker implantation, cardiac arrest, and vascular complications compared to SAVR. ^6^ However, this study did not assess the interaction with time and the type of AVR, thus, we are unable to deduce the association between AVR and changes in complications over time. Our study adds to existing evidence by including a more comprehensive list of complications, comparing the risk of complications over a long period of time for TAVR and SAVR, and presents respective temporal patterns for each (TAVR and SAVR).

Several caveats of the present analysis warrant consideration. The study was limited to patients enrolled in Medicare FFS, thus we were unable to study this association for the increasing number of Medicare Advantage patients. The retrospective cohort design of the present study relies on coding and Medicare FFS File claims and therefore many clinical parameters were ascertained through sources of automated data coding. Consequently, inherent to any largescale national audit of clinical practice relying on coding, this could have introduced bias in terms of over- or under-coding. We could only control for measured confounders, which is a limitation when examining non-fatal complications that may be influenced by anatomic factors, laboratory findings, and differences in comorbidity severity not captured in claims data. Nevertheless, a strength of the present analysis is that the data reflects real-world patient characteristics and outcomes for Medicare FFS patients across the country from different hospitals and physicians as compared to evidence from controlled clinical trials where many such patients are often excluded. Additionally, our temporal analysis ended in 2019, as we chose not to focus on the impact of different treatment patterns and a different healthcare environment during the COVID-19 pandemic on complication rates following AVR; additional research is needed on that topic.

In conclusion, in this large-scale study of temporal changes in overall in-hospital complications in patients treated with AVR across the US from 2012 through 2019, we demonstrate that overall complications rates have decreased, 4.5-fold greater for TAVR than SAVR. Additionally, greater numbers of patients overall are undergoing AVR, driven largely by the exponential growth in TAVR. These data, offering a snapshot of AVR-practice across the US from 2012-2019, offer important insights into the evolving nature of AVR, specifically the constantly improving procedural aspects, device improvements, and overall maturity of the procedure, especially TAVR.

## Data Availability

This retrospective cohort study used Medicare fee for service (FFS) data accessed through a license agreement and is not able to be posted to a website.

## Supplemental materials

### List of Figures

**Figure S1 (A-M):** Percentage of individual complications over time (unadjusted) in the elective isolated AVR cohort.

### List of Tables

**Table S1:** Multivariable model assessing the risk of complications in the elective isolated AVR cohort (TAVR vs SAVR over time)

**Table S2:** Multivariable Model Assessing Odds of Complications in the elective TAVR cohort over time

**Table S3:** Multivariable Model Assessing Odds of Complications in the elective isolated SAVR cohort over time

**Table S4:** Code list for complications included in the outcome of this study

